# Rare protein-disrupting variants in *CETP* and protection against vascular events

**DOI:** 10.1101/2024.12.02.24318306

**Authors:** Fredrik Landfors, Mathijs de Kleer, John J.P. Kastelein, Elin Chorell

**Affiliations:** Department of Public Health and Clinical Medicine, Section of Medicine, Umea University, S-901 87 Umea, Sweden; NewAmsterdam Pharma, Naarden, the Netherlands; Department of Vascular Medicine, Academic Medical Center, University of Amsterdam, the Netherlands

## Abstract

**Background:** Sequence variants that disrupt Cholesteryl Ester Transfer Protein (CETP) provide a genetic proxy for therapeutic inhibition, but their association with atherosclerotic vascular events has not been robustly established.

**Objectives:** To assess whether protein-disrupting *CETP* variants conferring lifelong reduced CETP activity lower atherosclerotic vascular event rates.

**Methods:** We tested associations between protein-disrupting *CETP* variants and atherosclerotic vascular and coronary artery disease across the All of Us Research Program and the UK Biobank using whole-genome sequences and linked health data. Predicted protein-disrupting variants were identified with LOFTEE and AlphaMissense. Gene-based burden testing used Firth’s logistic regression via REGENIE. Coronary artery disease was meta-analysed with FinnGen, Biobank Japan, and previous case-control studies. Restricted mean event-free survival to average life expectancy of 81 years was estimated in the UK Biobank.

**Results:** We included 1,515–4,575 carriers of *CETP* protein-disrupting variants and 848,000–1,585,000 non-carriers in combined populations with 126,000 atherosclerotic events and 246,000 coronary artery disease events, respectively. HDL cholesterol was 25% higher (0.35 mmol/L [14 mg/dL], *P*=4×10^-138^) and non-HDL cholesterol was 6% lower (0.23 mmol/L [9 mg/dL], *P*=9×10^-8^) in carriers. Atherosclerotic vascular event rates were lower in carriers (odds ratio, 0.73, 95% confidence interval, 0.62–0.87, *P*=3×10^-4^). Coronary artery disease risk was also lower in carriers (odds ratio, 0.79, 95% confidence interval, 0.72–0.86, *P*=3×10^-7^), reaching exome-wide significance. Carriers lived an average 1.0 years longer without atherosclerotic disease (95% confidence interval, 0.5-1.4, *P*=6×10^-5^).

**Conclusion:** Protein-disrupting *CETP* variants were associated with reduced atherosclerotic vascular disease risk and longer disease-free survival.

## Introduction

Cholesteryl ester transfer protein (CETP) is a lipid exchange protein that facilitates the transfer of cholesteryl esters from high-density lipoprotein (HDL) to low-density (LDL) or very-low-density lipoprotein (VLDL) in exchange for triglycerides.^1,2^ Because this lowers HDL cholesterol and raises atherogenic lipoproteins, CETP became an attractive therapeutic target, leading to the development of several small-molecule inhibitors. Clinical trials of these inhibitors, however, yielded mixed results. Torcetrapib, the first inhibitor, increased cardiovascular events and mortality through mineralocorticoid receptor activation, which increased aldosterone levels and blood pressure.^3,4^ Subsequent inhibitors, dalcetrapib and evacetrapib, failed to reduce cardiovascular events in outcomes trials.^5,6^ Anacetrapib showed promise by reducing vascular events in a phase 3 trial, but its development was halted due to its modest effect size, as well as accumulation in adipose tissue raising long-term safety concerns.^7,8^ Obicetrapib is in ongoing cardiovascular outcomes trials, with no oj-target effects reported to date.^9^

The failure of three of four major trials has cast doubt on the therapeutic value of CETP inhibition in atherosclerosis prevention, but whether the failures reflected poor pharmacological properties of the molecules tested or limitations intrinsic to CETP inhibition remains to be answered. Human genetics offers a way to address this question by isolating the consequences of reduced CETP activity from compound-specific oj-target effects. Heterozygous carriers of *CETP* protein-disrupting variants experience approximately one-third reduced CETP activity,^10,11^ effectively serving as natural experiments of lifelong, selective inhibition. Because genetic variants are assigned through random-like processes independent of environmental exposures occurring later in life, genetic studies are less susceptible to confounding and reverse causation, and can inform drug target assessment.^12–14^

Previous genome-wide association and Mendelian randomization studies have established robust associations between common variants at the *CETP* locus and both lipid levels and cardiovascular risk.^15–19^ Rare protein-disrupting variants offer a complementary approach. By producing more substantial, mechanistically interpretable reductions in protein quantity or function, they may more directly model the anticipated effects of pharmacological inhibition.^20^ Nomura et al. previously examined rare *CETP* protein-truncating variants and coronary heart disease.^21^ However, because protein-disrupting variant carriers are rare, prior estimates have been imprecise and have not reached exome-wide significance.

We therefore sought to confirm these findings with greater precision using larger biobanks. To do this, we ascertained *CETP* protein-disrupting variants in the UK Biobank and the All of Us Research Program to measure their association with incident and prevalent atherosclerotic vascular events, coronary artery disease, and plasma lipids. Analysing these two independent biobanks enabled replication with harmonized variant annotation and comparable outcome definitions in well phenotyped populations. We then conducted a meta-analysis combining our results with Biobank Japan, FinnGen, and the prior case-control study,^21^ to improve power and test consistency across cohorts and ancestries.

## Methods

### Study cohorts and outcome definitions

The UK Biobank (UKB) is a population-based study with health outcomes data, genetics, and blood chemistry from 500,000 participants across the United Kingdom.^22^ The study enrolled individuals aged 40-69 between 2006 and 2010, during which blood samples were collected. Ongoing follow-up ensures that health information is continuously updated through medical records, death certificates, and national registries. The data (UKB v20) were last updated on December 14, 2025. Atherosclerotic vascular disease events were defined as the first-occurring death certificate or hospital inpatient record diagnosis of coronary artery disease, ischemic stroke, peripheral atherosclerosis, precerebral atherosclerosis (carotid, vertebral, basilar), or abdominal aortic aneurysm. We also ascertained cases using procedure codes indicating revascularization for any atherosclerotic vascular disease endpoint. The international classification of disease (ICD) and OPCS Classification of Interventions and Procedures codes corresponding to each condition is found in **Supplemental Table 1**. LDL-C was measured both directly (UKB data field 30780), and calculated using the Friedewald formula from total cholesterol, HDL, and triglycerides (UKB data fields 30690, 30760, 30870). Non-HDL cholesterol was calculated by subtracting HDL from total cholesterol. Apolipoprotein A-I, B, and Lipoprotein(a) levels were measured using immunoturbidimetry (UKB data fields 30630, 30640, 30790).

The All of Us Research Program is a longitudinal cohort study launched by the National Institutes of Health in 2018, having enrolled more than 873,000 participants as of 2026.^23^ Participants contribute with their electronic health records, complete surveys, provide biological samples, and undergo physical measurements. Currently, over 414,000 participants are included in whole-genome sequencing data set. The data (All of Us Controlled Tier Dataset v8) were last updated on January 13, 2026. Coronary artery disease, ischemic stroke, precerebral atherosclerosis, peripheral atherosclerosis, and abdominal aortic aneurysm were ascertained using Observational Medical Outcomes Partnership Common Data Model (OMOP CDM) codes (**Supplemental Table 2**). Total cholesterol, HDL, and triglycerides (OMOP concept IDs 3027114, 3007070, 3022192), were extracted from electronic health records, and LDL cholesterol was calculated from these values using the Friedewald formula. Non-HDL cholesterol was calculated by subtracting HDL from total cholesterol.

Prevalent coronary artery disease was previously ascertained in Biobank Japan, FinnGen, and one case-control study by Nomura et al.^21^ Biobank Japan recruited 200,000 individuals from 66 hospitals between 2003-2007. The case definition was being positive for myocardial infarction (ICD-10: I21/I22/I23/I24/I25), unstable angina (ICD-10: I20.0), or stable angina (ICD-10: I20.9) diagnoses.^24,25^ FinnGen consists of a network of biobanks that integrates genetics with longitudinal health data from national registries for over 520,000 Finns.^26^ Coronary artery disease cases had a hospital discharge record, insurance reimbursement or death certificate positive for angina pectoris (ICD-10: I20), acute myocardial infarction or complications thereof (ICD-10: I21-I24), or chronic ischemic heart disease (ICD-10: I25), or corresponding ICD-8/9 and insurance codes (https://risteys.finngen.fi/endpoints/I9_IHD). Nomura et al. included 12 case-control studies with 67,281 participants.^21^ The included studies case definitions included predominantly early-onset myocardial infarction and early-onset coronary artery disease. To avoid sample overlap, the Biobank Japan subset (N=15,598) of Nomura et al. was excluded from the present meta-analysis.

### Pre-processing and quality control of genetic data

We used whole-genome sequences from approximately 460,000 UK Biobank and 390,000 All of Us participants. Whole-genome sequencing of the UK Biobank was performed between 2019 and 2022 using Illumina NovaSeq platforms, with data processed using DRAGEN v3.7.8 aligned to GRCh38 at 32.5x mean coverage.^27^ Whole-genome sequencing data from the All of Us Research Program were accessed via the Researcher Workbench Controlled Tier Dataset v8, processed with DRAGEN v3.7.8 at ≥30x mean coverage aligned to GRCh38 as described previously.^28^ Biobank Japan participants were genotyped using Illumina arrays and imputed using a combined 1000 Genomes and Japanese reference panel.^29^ FinnGen participants were genotyped using Illumina and Affymetrix arrays and imputed using a Finnish-specific reference panel.^26^ Nomura et al. participants underwent exome sequencing or genotyping.^21^

Genetic ancestry in UK Biobank was inferred using a support vector machine classifier implemented in KING,^30^ trained on 1000 Genomes reference populations and applied to principal components derived from genomic data. For All of Us, ancestry assignments provided by the program (v8 ancestry predictions) were used. We restricted analyses to ancestry groups where more than 5 participants with protein-disrupting variant were expected to experience the primary outcome event, using a carrier frequency of 2‰ and an event rate of 10% for calculations. Based on this criterion, participants of European ancestry were included from UK Biobank, and participants of African, Admixed American, and European ancestry were included from All of Us.

For REGENIE Step 1, we created ancestry-stratified sets of approximately 500,000 linkage disequilibrium-pruned variants. Variants were filtered based on minor allele frequency (MAF ≥1.5% for UK Biobank; ≥1% in All of Us), minor allele count (MAC ≥100 for UK Biobank; ≥20 for All of Us), Hardy-Weinberg equilibrium exact test (HWE, *P*>10^-15^), and genotype missingness (<10%). Linkage disequilibrium pruning was performed using a 500kb window and r^2^ threshold of 0.2. Individuals with >10% missing genotypes were excluded. For burden testing of the *CETP* locus (chr16:56,961,923-56,983,845; GRCh38) in REGENIE Step 2, variants were extracted from BGEN-formatted files with more permissive filtering (MAC≥1, HWE exact test *P*>10^-5^, genotype and sample missingness <10%) to include rare variants. Multiallelic variants were excluded in Step 1 but retained in Step 2.

For gene-based burden testing, we grouped variants based on predicted variant functional consequences. We restricted analyses to variants present in gnomAD v4.1.0 to ensure high-quality variant calls and enable consistent annotation across cohorts.^31^ Predicted protein-truncating variants (PTVs) included splice acceptor/donor, stop gained, and frameshift variants. High-confidence PTVs were defined as PTVs passing Loss-Of-Function Transcript Effect Estimator (LOFTEE) filters, which identify and exclude probable false-positive loss-of-function calls based on sequence context and splice predictions.^32^ Missense variant pathogenicity was predicted using AlphaMissense, a deep learning model based on AlphaFold that uses protein structure and evolutionary conservation to classify missense variants,^33^ with variants classified as “likely pathogenic” (score ≥0.564) included in the missense burden mask. For the gene-based burden tests, high-confidence PTVs were combined with AlphaMissense likely pathogenic variants into a single variable, designating their grouping as protein-disrupting variants.

### Statistical analysis

Association analyses were performed using REGENIE v4.1.^34^ REGENIE employs a two-step whole-genome regression approach. Step 1 fits a null model using a set of linkage disequilibrium-pruned common variants to account for population structure and relatedness through leave-one-chromosome-out predictions.^34,35^ Step 2 tests associations at individual variants or using gene-based burden masks while conditioning on the Step 1 predictions. Covariates included age, age^2^, sex, and the first 10 genomic principal components. For continuous traits, we used linear regression (--qt), and for binary outcomes, Firth-corrected logistic regression (--bt, --firth, --firth-se). Meta-analysis across ancestries and cohorts was performed using fixed-effects inverse variance-weighting implemented in the *metafor* R package.^36^ Study heterogeneity was estimated using Cochran’s Q test and calculating the I^2^ statistic. Exome-wide significance was defined as P<2.5×10^-6^ (0.05/20,000 genes), with uncorrected *P* values and 95% confidence intervals reported in **Figures 1-3**.

**Figure 1.**
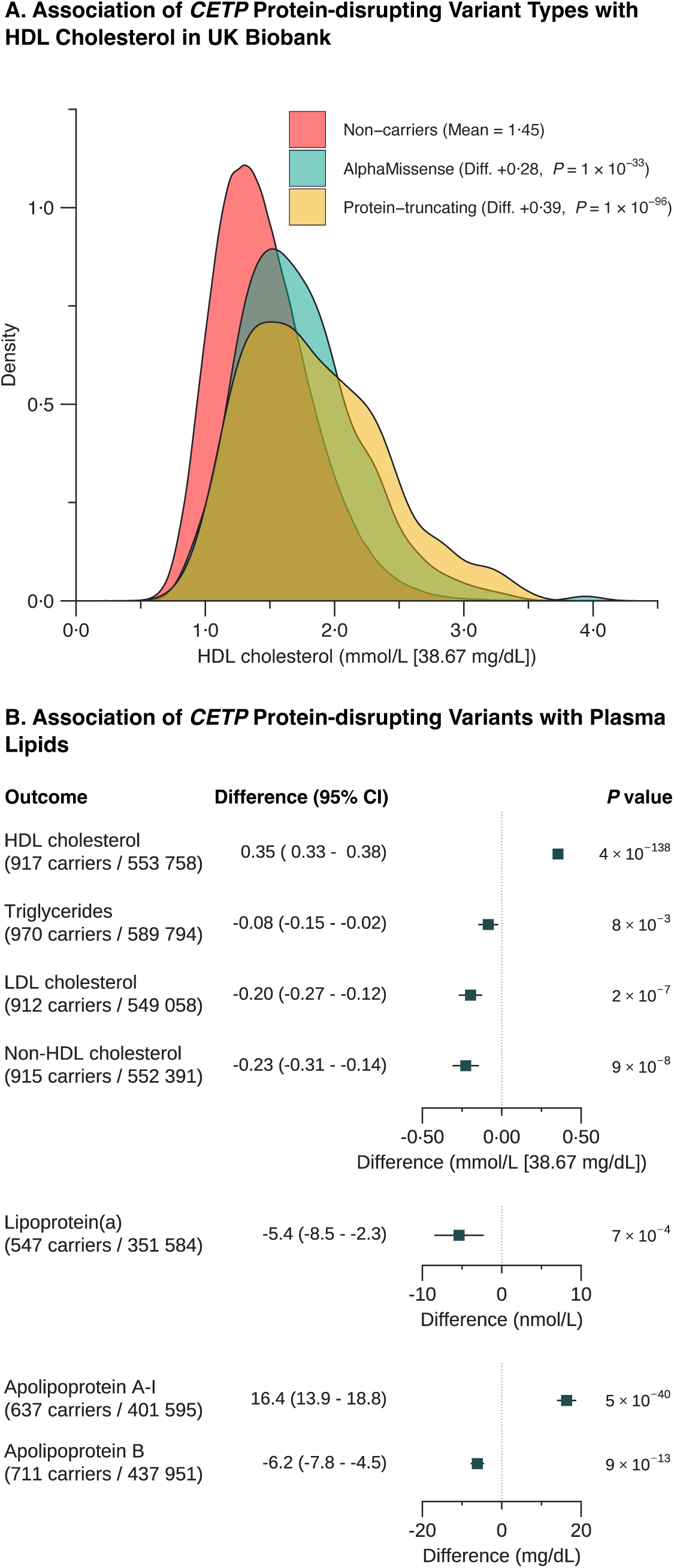
Association of *CETP* protein-disrupting variants with plasma lipids. **(A)** Distribution of HDL cholesterol concentrations in UK Biobank participants stratified by *CETP* variant status: non-carriers (red, mean 1.45 mmol/L [56 mg/dL]), AlphaMissense-predicted pathogenic variant carriers (blue; difference +0.28 mmol/L [11 mg/dL], P=1×10^-33^), and protein-truncating variant carriers (yellow; difference +0.39 mmol/L [15 mg/dL], P=1×10^-96^). **(B)** Forest plots showing the association between *CETP* protein-disrupting variant carrier status and plasma lipid concentrations in the combined UK Biobank and All of Us analysis adjusting for age, age^2^, sex, and the first 10 genomic principal components. Apolipoprotein A-I and apolipoprotein B were available in UK Biobank only. Error bars represent 95% confidence intervals.

**Figure 2.**
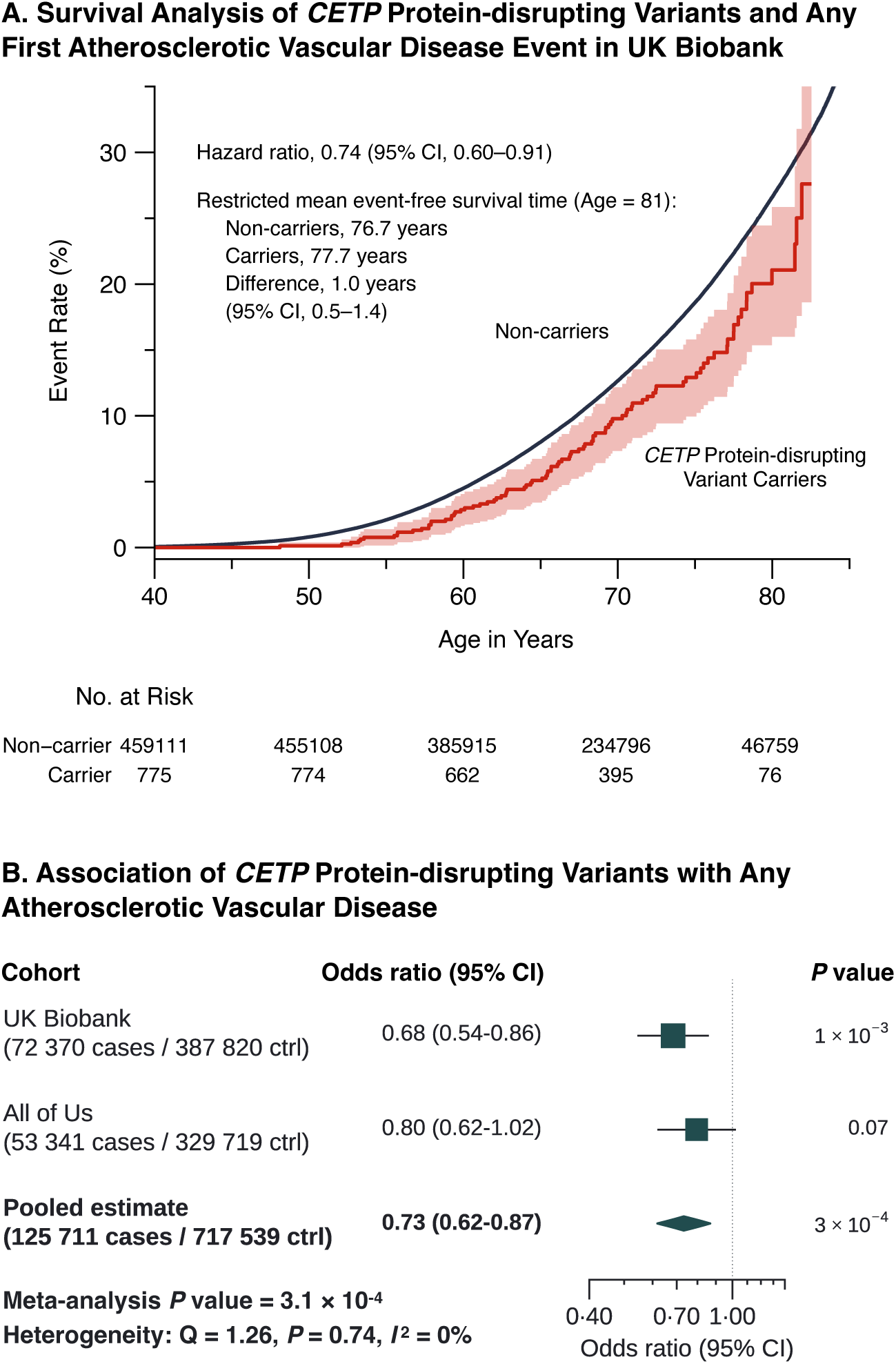
Association of *CETP* protein-disrupting variants with atherosclerotic vascular disease. **(A)** Cumulative incidence of first atherosclerotic vascular disease event by age among *CETP* protein-disrupting variant carriers and non-carriers in the UK Biobank. Shaded areas represent 95% confidence intervals. The crude unadjusted hazard ratio is shown. **(B)** Forest plot showing odds ratios for composite atherosclerotic vascular disease (coronary artery disease, ischemic stroke, peripheral atherosclerosis, precerebral atherosclerosis (carotid, vertebral, basilar), or abdominal aortic aneurysm) in *CETP* protein-disrupting variant carriers versus non-carriers adjusting for age, age^2^, sex, and the first 10 genomic principal components, stratified by cohort and genetically inferred ancestry. For All of Us, the pooled multi-ancestry estimate is presented to comply with data protection policies prohibiting reporting of results derived from cell counts <20. Heterogeneity statistics were calculated including UK Biobank European ancestry and All of Us Admixed American, African, and European ancestry substudies individually in the fixed-effects inverse variance-weighted estimator.

**Figure 3.**
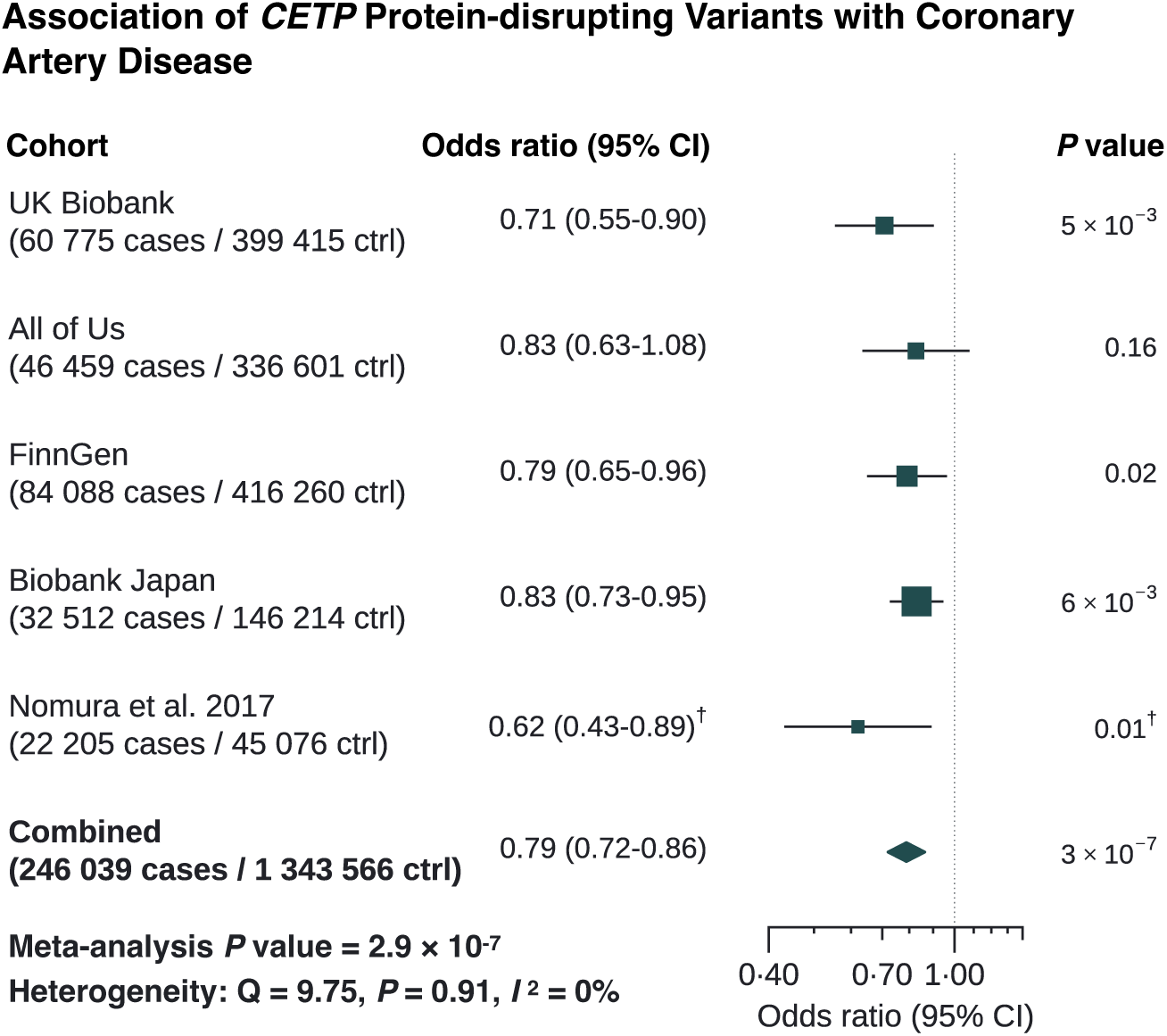
Meta-analysis of *CETP* protein-disrupting variants and coronary artery disease. Forest plot showing the association between *CETP* protein-disrupting variant carrier status and coronary artery disease across UK Biobank, All of Us, FinnGen, Biobank Japan, and case-control studies from Nomura et al. (2017).^21^ The pooled multi-ancestry estimate is displayed for All of Us to comply with data protection policies that prohibit reporting summary statistics derived from cell counts <20. For consistency, Nomura et al. (2017) is also displayed as a combined estimate. Heterogeneity statistics were calculated using individual substudies from All of Us (n=3) and Nomura et al. (n=12). ^†^ The Nomura et al. (2017) combined estimate excludes the Biobank Japan substudy (N=15,598) to prevent sample overlap, and therefore differs from the originally reported estimate.^21^

In the UK Biobank, time-to-event analyses were conducted to estimate the cumulative incidence of atherosclerotic vascular disease by carrier status. Exposure to a defective *CETP* allele was assumed to begin after birth, and participants were followed from their birth (UKB data fields 34, 54) until the occurrence of the first event, lost to follow-up (UKB data field 191), death (UKB data field 40000), or the last date registries were queried (Scotland, death registry 2024-11-30, hospital inpatient 2022-08-31; England, death registry, 2024-08-31, hospital inpatient 2023-03-31; Wales, death registry 2024-08-31, hospital inpatient 2022-05-31), whichever occurred first. Kaplan-Meier survival curves were constructed, and the crude hazard ratio was measured using Cox regression. To estimate the lifetime benefit, restricted mean survival time was calculated using the *survRM2* R package.^37^ The restriction time of 81 years was set based on the U.K. average life expectancy for 2023.^38^

### Ethical Review

This research has been conducted using the UK Biobank Resource under Application Number 148828. All of Us data were accessed under an institutional data use agreement established the All of Us Research Program and the analyses were approved by the Swedish Ethical Review Authority (Dnr 2025-05711-01). Summary statistics from Biobank Japan, FinnGen, and Nomura et al. are publicly available,^21,24,26^ and analyses of these data did not require separate ethical approval. All participants provided written informed consent, and the research adhered to the ethical standards set forth in the Declaration of Helsinki.

## Results

### Cohort characteristics and protein-disrupting variants

Descriptive characteristics on the included 849,761 UK Biobank and All of Us participants are presented in **Table 1**. The mean age at recruitment into the UK Biobank was 56.8 years and 45.8% were male. The mean age of recruitment into the All of Us cohort was 58.0 years and 39.0% were male. We identified 775 *CETP* protein-disrupting variant carriers in the UK Biobank, whereof 463 (59.7%) were LOFTEE-predicted high-confidence protein-truncating variants and 312 (40.3%) were AlphaMissense-predicted pathogenic variants. In All of Us, 740 protein-disrupting variant carriers were identified, whereof 437 (59.1%) participants carried high-confidence protein-truncating variants, and 303 (40.9%) participants carried AlphaMissense-predicted pathogenic variants.

**Table 1.**
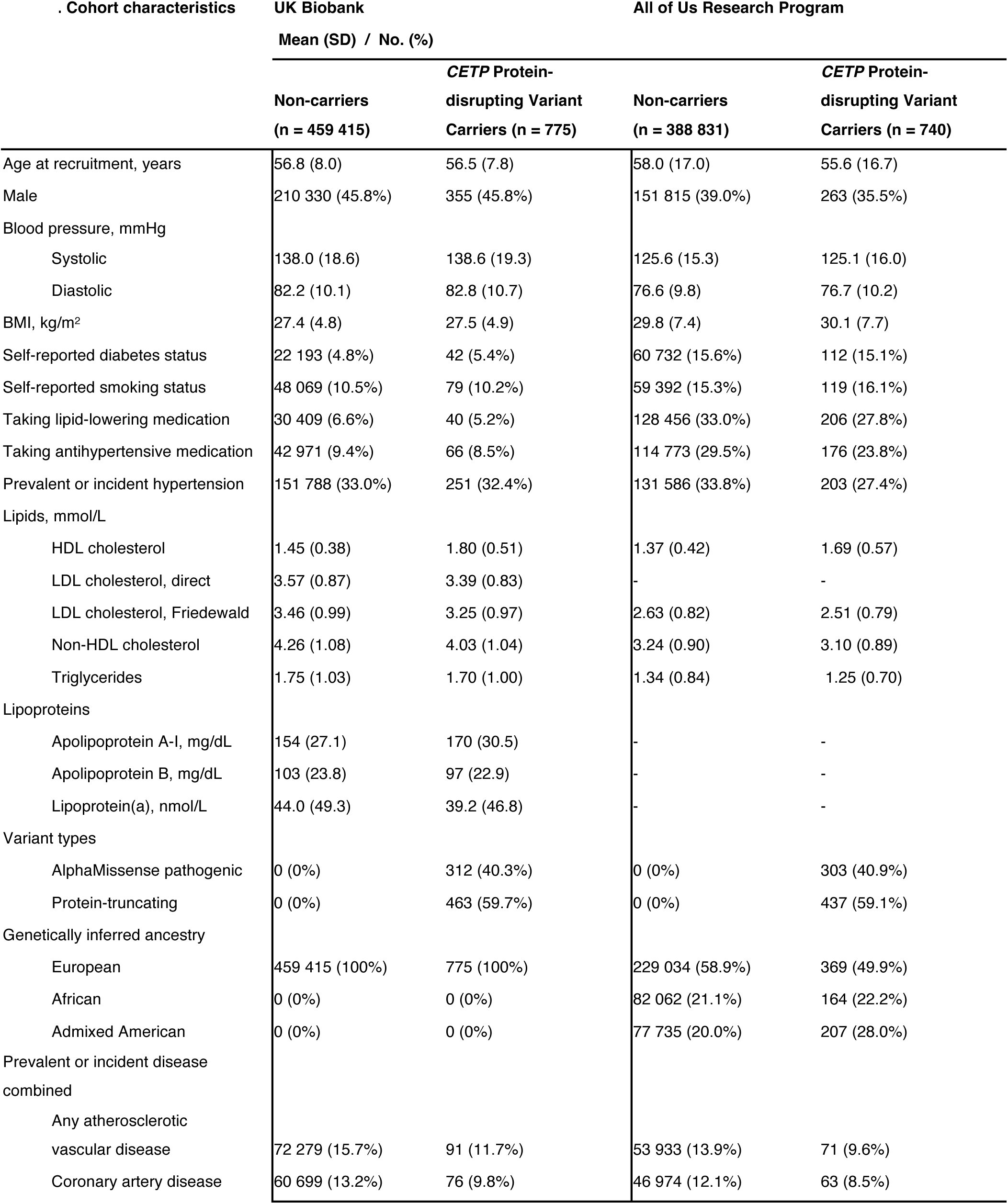
Cohort characteristics of the included UK Biobank and All of Us participants.

To confirm that both AlphaMissense and LOFTEE-predicted protein-disrupting variants altered plasma CETP activity, we measured their association with HDL cholesterol and plotted groupwise HDL cholesterol distributions in the UK Biobank. We tested each variant with minor allele count ≥5 individually for their association with HDL cholesterol. One relatively common AlphaMissense-classified pathogenic variant (GRCh38 chr16:56,973,332:G>T [p.Gly251Val], European ancestry MAF 0.19‰, HWE-estimated N_carriers_=188), showed no association with HDL cholesterol (fixed effects estimate, +0.0069 mmol/L [0.3 mg/dL], 99.99975% CI: −0.133 to 0.15, *P*=0.82) across All of Us and UK Biobank, reliably excluding the expected effect size for *CETP* protein-disrupting variants (≥0.20 mmol/L [8 mg/dL]). This variant was excluded from gene-based burden testing. All other predicted *CETP* protein-disrupting variants identified in UK Biobank and All of Us were included in the gene-based burden tests and are listed in **Supplemental tables 3-4**. To further ensure variants included in burden testing disrupted CETP function, we plotted their distribution in UK Biobank. Both AlphaMissense-predicted pathogenic variants and LOFTEE-predicted high-confidence protein-truncating variants raised mean HDL cholesterol by 20% (+0.28 mmol/L [11 mg/dL], *P*=1×10^-33^) and 27% (+0.39 mmol/L [15 mg/dL], *P*=1×10^-96^) respectively, with both groups showing concordant rightward-shifted distributions relative to non-carriers (**Figure 1A**). These effect estimates were within the expected range of known *CETP* variants that cause complete CETP deficiency in their homozygous state.^10^

### Association with lipids and atherosclerotic vascular disease

The associations between *CETP* protein-disrupting variant carrier status and plasma lipids are shown in **Figure 1B**. In the combined All of Us and UK Biobank analysis, *CETP* protein-disrupting variant carrier status was associated with 25% higher HDL cholesterol, at +0.35 mmol/L (14 mg/dL, *P*=4×10^-138^). Non-HDL cholesterol was 6% lower at −0.23 mmol/L (9 mg/dL, *P*=9×10^-8^), and Friedewald LDL cholesterol was 6% lower at −0.20 mmol/L (8 mg/dL, *P*=2×10^-7^). In UK Biobank, an exome-wide significant 6% reduction in apolipoprotein B concentration (*P*=9×10^-13^) was also observed, along with an 11% increase in apolipoprotein A-I (*P*=5×10^-40^), and a nominal 12% reduction in lipoprotein(a) (*P*=7×10^-4^).

The association of *CETP* protein-disrupting variant carrier status with composite atherosclerotic vascular disease in the UK Biobank and All of Us cohorts is shown in **Figure 2**. Prevalent and incident atherosclerotic vascular disease, defined as coronary artery disease, ischemic stroke, peripheral atherosclerosis, precerebral atherosclerosis, or abdominal aortic aneurysm, was ascertained in 72,370 of 460,190 (16.1%) UK Biobank participants and in 53,341 of 383,060 (13.9%) All of Us participants. In fixed-effects meta-analysis combining estimates from UK Biobank with All of Us, carriers had 27% lower odds of atherosclerotic vascular disease (odds ratio, 0.73, 95% CI: 0.62-0.87, *P*=3×10^-4^). Time-to-event analyses in the UK Biobank showed *CETP* protein-disrupting variant carriers lived an average of 1.0 years longer without atherosclerotic disease compared to non-carriers (95% CI: 0.5-1.4, *P*=6×10^-5^), with mean survival times estimated at 77.7 years and 76.7 years, respectively.

### Coronary artery disease meta-analysis

The association between *CETP* protein-disrupting variant carrier status and coronary artery disease in 246,000 cases and 1,344,000 controls is shown in **Figure 3**. Meta-analysis was conducted by including Biobank Japan (32,512 cases/146,214 controls; mean age 63 years at donation, 54% male),^24^ FinnGen (84,088 cases/416,250 controls; mean age 53 years at donation, 44% male),^26^ and previous case-control studies (22,205 cases/45,076 controls; mean age 52 years, 57% male) published by Nomura et al.,^21^ together with the UK Biobank and All of Us. Since the broad atherosclerotic vascular disease composite used in the previous analyses were unavailable in Biobank Japan, FinnGen and the 12 case-control studies, the association between coronary artery disease and *CETP* protein-disrupting variant carriers was measured in All of Us (46,459 cases/336,601 controls) and the UK Biobank (60,775 cases/399,415 controls) to ensure harmonized endpoints.

In Biobank Japan, the East Asian-enriched (88-fold vs. non-Finnish Europeans) 16:56,982,238:G>A (GRCh38; c.1321+1G>A) splice donor LOFTEE-predicted high-confidence PTV was present in approximately 2,008 participants (estimated from HWE equations based on MAF 5.63‰), well-imputed (INFO=0.93), and associated with raised HDL cholesterol (+0.32 mmol/L [12 mg/dL], *P*=5×10^-122^) and lowered LDL cholesterol (−0.16 mmol/L [6 mg/dL], *P*=5×10^-7^). The HDL cholesterol-raising effect was well within the expected range of protein-disrupting *CETP* variants in their heterozygous state, and homozygous carrier status of this variant is known to cause complete CETP deficiency with extremely elevated HDL cholesterol concentrations (mean 4.2 mmol/L [162 mg/dL] vs. 1.4 mmol/L [54 mg/dL] in unaffected family members).^10^ The Finnish-enriched (48-fold vs. non-Finnish Europeans) splice donor variant 16:56,962,097:TGTAA>T (GRCh38; c.118+4_118+7del) was well-imputed (median INFO=1.00), and present in approximately 895 participants. Association analysis in the UK Biobank found this variant was nominally associated with raised HDL cholesterol (+0.22 mmol/L [9 mg/dL], *P*=0.008) and directionally consistent but non-significantly associated with lowered non-HDL cholesterol (−0.46 mmol/L [18 mg/dL], *P*=0.07). Nomura et al. found 279 splice acceptor/donor, stop gained, and frameshift variant carriers with raised HDL cholesterol (+0.58 mmol/L [22 mg/dL], *P*=1×10^-4^), and lowered LDL cholesterol (−0.31 mmol/L [12 mg/dL], *P*=0.043).^21^ Because one Nomura et al. substudy (N=15,598) overlapped with Biobank Japan, substudies including 157 carriers were included in the present meta-analysis.

Fixed effects inverse variance-weighted meta-analysis found the risk of coronary artery disease was lower in *CETP* protein-disrupting variant carriers (odds ratio, 0.79, 95% CI: 0.72-0.86, *P*=3×10^-7^), meeting the gene-based exome-wide significance threshold. Effect estimates were directionally consistent across the contributing studies, with no significant heterogeneity (I^2^=0%).

## Discussion

In this multi-ancestry analysis of up to 1,586,000 participants including 4,575 carriers of *CETP* protein-disrupting variants, we observed a 21% reduction in coronary artery disease odds that reached exome-wide significance, along with significant reductions in composite atherosclerotic vascular disease. Carriers of *CETP* protein-disrupting variants lived an average of one additional year without atherosclerotic vascular disease compared to non-carriers in the UK Biobank. Together, these findings provide robust and convergent human genetic evidence that *CETP* loss-of-function is associated with reduced atherosclerotic disease risk.

Several analyses of common *CETP* variants have shown that genetically reduced CETP activity lowers cardiovascular risk.^15–19^ Rare protein-disrupting variants offer complementary evidence. They typically produce substantial reductions in CETP activity through a less ambiguous loss-of-function mechanism, more directly modelling the effects anticipated with pharmacological inhibition. Consistent with this, these variants were associated with higher apolipoprotein A-I and HDL cholesterol as well as lower LDL cholesterol, apolipoprotein B, and lipoprotein(a), mirroring the effects on lipids observed with pharmacological CETP inhibition in clinical trials.^39^ Extending the analysis by Nomura et al.,^21^ our larger study achieved gene-based exome-wide significance for the association between *CETP* protein-disrupting variants and coronary artery disease (*P*=3×10^-7^), strengthening the evidence that *CETP* loss-of-function lower the risk of atherosclerotic vascular events.

These results align with principles for using human genetics to evaluate drug targets.^20^ The genetic variants examined were predicted to disrupt CETP function through truncation or protein-damaging missense substitutions, providing a link between genotype and reduced plasma CETP activity. Protein-truncating variants typically act through nonsense-mediated mRNA decay, though variants escaping mRNA decay may produce truncated proteins that are unstable or non-functional,^40^ while AlphaMissense-predicted pathogenic missense variants may reduce protein abundance through destabilization and degradation, or impair function directly by substituting catalytically or structurally critical residues.^33^ The similar effects of both variant classes on HDL cholesterol distributions, as shown in **Figure1A**, support their inclusion in a unified burden test.

Several limitations warrant consideration. First, the variants we studied represent constitutive germline effects that act systemically, from development through adulthood, in all *CETP*-expressing tissues, whereas clinical trials administer drugs with different tissue distribution and target selectivity, to adults, and generally follow participants for only a few years. Lifelong genetic effects may confer benefits such as prevention of early atherosclerotic plaque formation, that are not achievable with pharmacological intervention later in life. Conversely, the cumulative years of exposure in genetic variant carriers may exceed what is practically achievable in a clinical trial setting, which could inflate estimates when translating genetic effects onto what effect would be achievable in trials. Second, we examined heterozygous carriers who exhibited approximately 25% higher HDL cholesterol levels (+0.35 mmol/L [9 mg/dL]), and 6% lower LDL cholesterol levels (−0.20 mmol/L [8 mg/dL]). This effect magnitude is consistent with earlier observations that partially CETP-deficient heterozygotes typically show 25% increases in HDL cholesterol,^10,41,42^ and retain 58–68% of normal CETP activity, suggesting partial compensation by the wild-type allele.^11^ However, this one-third inhibition only represents approximately one-fifth to one-seventh of the lipid-altering potency of pharmacological CETP inhibitors, which can raise HDL cholesterol by up to 140% and lower LDL cholesterol by up to 44% at near-complete CETP inhibition in participants without background lipid-lowering therapy.^43^ Investigation of homozygous carriers, who exhibit complete CETP deficiency, is challenging and was not feasible due to their extreme rarity. The cardiovascular benefits of the near-complete CETP inhibition achieved with potent CETP inhibitors therefore require confirmation in clinical trials. Third, FinnGen and Biobank Japan genotypes were imputed from reference panels rather than directly sequenced, albeit at high quality (INFO=0.93–1.00). Incorrect imputation of rare variants typically results in genotypes homozygous for the reference allele, producing one-sided misclassification that biases toward the null.^44^ The FinnGen and Biobank Japan estimates are therefore likely conservative rather than inflated by imputation error. Fourth, restricted mean survival time estimates reflect the baseline risk of the study populations and may differ in populations with higher or lower cardiovascular risk. Fifth, the inferential validity of genetic association studies relies on the assumption that genetic variants are distributed in a random-like pattern within the study population, mimicking randomization in a clinical trial. Potential confounders such as population stratification or assortative mating could bias estimates by creating non-causal correlations between *CETP* variants and cardiovascular events.^45^ We sought to minimize these biases by stratifying analyses by genetically inferred ancestry groups, adjusting for genomic principal components within those groups, and demonstrating consistent effect estimates across multiple independent cohorts and ancestries, which together mitigate concerns about confounding.

In conclusion, rare protein-disrupting variants in *CETP* conferring lifelong inhibition were associated with reduced atherosclerotic vascular disease risk and longer atherosclerotic vascular disease-free survival. These findings add human genetic evidence for the role of CETP inhibition in atherosclerotic vascular disease prevention and supports CETP as a therapeutically relevant target. Outcomes trials will determine the role of small-molecule CETP inhibitors in cardiovascular prevention.

## Acknowledgements

We gratefully acknowledge All of Us participants for their contributions, without whom this research would not have been possible. We also thank the National Institutes of Health’s All of Us Research Program for making available the participant data examined in this study. We want to acknowledge the participants and investigators of the FinnGen study. We thank the participants and hospital group of Biobank Japan. This research has been conducted using the UK Biobank Resource under Application Number 148828. This work uses data provided by patients and collected by the NHS as part of their care and support.

## Data availability

UK Biobank data are available to authorized researchers through application (https://www.ukbiobank.ac.uk). This study used data from the All of Us Research Program’s Controlled Tier Dataset v8, available to authorized users on the Researcher Workbench (https://www.researchallofus.org). FinnGen summary statistics are publicly available (https://www.finngen.fi/en/access_results). Biobank Japan summary statistics are publicly available (https://pheweb.jp). The case-control data from Nomura et al. were obtained from the published manuscript.^21^ The UK Biobank data field identifiers and OMOP concept IDs used in the analyses are described in the **Methods** section and **Supplemental Tables 1-2**.

## Appendices

### Supplemental tables

*Supplemental table 1*

**UK Biobank disease definitions**

Disease phenotype definitions used for outcome ascertainment in UK Biobank.

*Supplemental table 2*

**All of Us disease definitions**

Disease phenotype definitions used for outcome ascertainment in the All of Us Research Program.

*Supplemental table 3*

**UK Biobank *CETP* protein-disrupting variants.**

*CETP* variants classified as protein-disrupting identified in UK Biobank whole-genome sequencing data.

*Supplemental table 4*

**All of Us *CETP* protein-disrupting variants.**

*CETP* variants classified as protein-disrupting identified in All of Us Research Program.

